# Signatures of medications in meditation: A Connectivity map analysis of transcriptomes from an inner engineering retreat

**DOI:** 10.1101/2023.11.27.23299046

**Authors:** Aditi Joshi, Deep Patel, Mitali Mukerji

**Author notes:** Equal contribution (Aditi Joshi, Deep Patel). MM conceptualised and designed the study, DP, AJ and MM carried out the analysis, AJ and MM wrote the manuscript.

## Abstract

Meditation practices, known for their stress management and well-being benefits, are being increasingly integrated into wellness regimens and adjunct therapies for chronic conditions. We propose that beneath their non-pharmacological facade, meditation practices might operate via drug target modulation. Here, we leverage the Connectivity Map (CMap) to investigate (a) the overlap between meditation-induced molecular signatures and established drug responses, and (b) the pathways and mechanisms contributing to meditation therapeutic effects. This was studied in a comprehensive temporal RNAseq dataset comprising premeditation, meditation, and follow-up stages from a clinical trial involving 106 participants practising inner engineering meditation. Most striking, we observed intersection of meditation signatures with over 438 drugs, with ≥98% negative and positive connectivity scores and clusters of individuals with differential response. These drugs predominantly target the neuroactive ligand receptor signaling pathway, that are used widely in neuro-psychiatric disorders, hypertension, migraine, pain, insomnia, nicotine addiction, alcoholism, and cancer. This study underscores the need to (a) approach meditation with the same caution as medication, (b) tailor and calibrate meditation practices based on individual health status, disease profile, and concurrent medications, and (c) conduct meditation under expert guidance.

---

Meditation methods, which have their roots in Indian medicine and Buddhism, are used as non-pharmacological adjunct therapies for managing stress, sleeplessness, anxiety, pain, depression, and cognitive improvement(1). Meditation promotes rest, relaxation, and digestion by stimulating the parasympathetic nerve system, which counteracts the sympathetic “fight-or-flight” response(2). However, the physiological effects differ between individuals and are altered by procedures and durations, possibly resulting in a range of outcomes(3)(4). Meditation, might aggravate symptoms in people who already have mental health issues. A recent transcriptome study on advanced inner engineering meditation reported modulations of networks that are perturbed in severe COVID-19 infection and multiple sclerosis. This suggests meditation may be useful in conditions of compromised immune systems and increased inflammation(5). We hypothesized that the molecular targets underlying meditation’s response might overlap with those modulated by pharmacological interventions. We used connectivity map (CMap) to query intersections of meditation response with over 3 million gene expressions of 5,000 small compounds and 3,000 genetic perturbations across 9 cell lines(6)(7). Our study revealed convergence with over 438 pharmaceutical drugs, which were strongly connected to neuroactive ligand receptor signalling pathways. This study emphasizes that similar to medications there is a need for a cautious approach to the seemingly non-pharmacological practice of meditation. This should be carried out under the supervision of knowledgeable practitioners to ensure optimal and customized results.

## 1. Results

We leveraged the demonstrated capabilities of the connectivity map, to discern the potential pharmacological connections during meditation(7). This was tested using RNAseq-based transcriptomes from a clinical study involving 106 participants in an advanced inner engineering meditation retreat program(5). The time course data, spanning from the initial phase (T1) through the 8-week preparatory phase (T2), the 8-day inner engineering retreat (T3), and a 3-month follow-up (T4), enabled us to explore specific effects.

### A. Connectivity Analysis of Meditation Transcriptome reveals Potential Drug Targets

CMap revealed a significant intersection with over 438 drug signatures across the three time spans (Fig:1). There was a significant overlap in profiles of drug signatures in the preparatory and the meditation phase (T1vsT2, T2vsT3) which was were very distinct from the follow-up phase (T2vsT4). The drug classes identified were diverse, encompassing cyclooxygenase inhibitors, serotonin receptor agonists/antagonists, glucocorticoid receptor agonists, dopamine receptor antagonists, HDAC inhibitors to name a prominent few (Table:S1). Gene ontology (GO) analysis of the drug targets (a) drugs with positive scores and knockdown genes and (b) negative scores and over-expressed genes highlighted the enriched pathways through which meditation could exhibit its effects (Fig:1). Noteworthy, the neuroactive ligand receptor interaction pathway (hsa04080) consistently featured within the positive connectivity set across all three phases of meditation. The targets were quite distinct in the follow-up phase and corroborates with the patterns from drug signatures (Fig:1,Fig:S1).

**Fig. 1.**
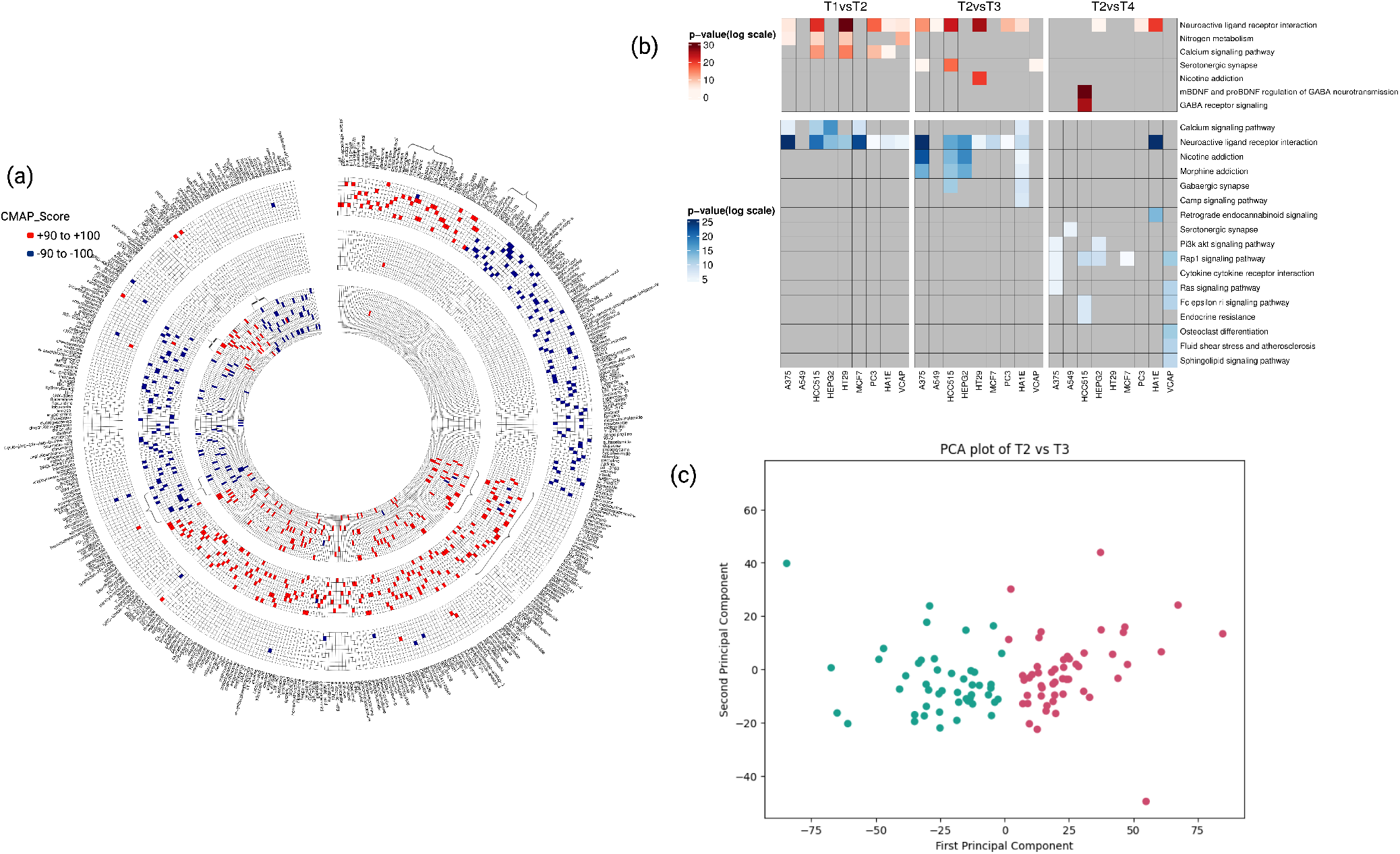
(a) Circos Plot of Connectivity Map Matching Signatures .The Circos plot depicts the Connectivity map analysis results for matching signatures of drugs across three distinct time points within subjects who participated in the advanced inner engineering meditation retreat. The time points under consideration are: before the retreat (T1), after an 8-week preparatory phase (T2), 8 days after the Samyama meditation phase (T3), and 3 months after the retreat (T4). Each concentric circle represents a specific comparison: the innermost circle corresponds to the connectivity of drugs with differential expression signatures from T1 vs T2, the middle circle represents T2 vs T3, and the outermost circle corresponds to T2 vs T4. In the visualization, drugs exhibiting connectivity with positive scores equal to or exceeding 98 are color-coded in red, while those with negative scores are depicted in blue. Remarkably, there is a substantial overlap of drug signatures between the preparatory and meditation phases. Notably, the number of drugs displaying connectivity in T2 vs T3 (243) surpasses that of the preparatory phase (210). Interestingly, after 3 months, a majority of these initial signatures dissipate, giving way to the emergence of new drug signatures. The drugs targeting the significantly enriched neuroactive ligand receptor signaling pathway are denoted by the symbol.(b) Pathway enrichment analysis of proteins targeted by drugs across all three phases. The upper panel, highlighted in blue, showcases significant pathway enrichments based on Log p values derived from drugs with positive scores. The lower panel depicts enrichments from drugs with negative scores.(c) The PCA plot of sample wise expression difference in T2 vs T3 reveals two distinct clusters of meditation response within the study cohort. Image created with BioRender.com

### B. Inter-individual variability in meditation response

Intersection with such a large number of drugs could be a cumulative effect of differential response of individual samples. To test this, we carried out a sample-wise TMM normalization followed by differential expression analysis between T2 and T3 (P *<* 0.05), PCA and hierarchical clustering. Two distinct clusters of samples separated out in PCA that had distinct non-overlapping connectivity with drugs (Fig:1, Fig:S1).

## 2. Discussion

The aim of this study was to explore the potential connections between the mechanisms underpinning non-pharmacological meditation techniques and medication targets. A connectivity map study of transcriptomes from 106 participants in an inner engineering meditation retreat revealed strong connections with over 438 medicines used in neuro-psychiatric, cancer, addiction, and hypertension therapies. The clinical study(3)(8) had also revealed a number of positive outcomes, including altered levels of cellular anandamide, antiinflammatory and analgesic reactions, vascular relaxation, and decreased levels of atherosclerosis-associated metabolites that correlate with the drug signatures. Many of the drugs obtained through CMap target GABA, endocannabinoid, dopamine, serotonin, acetylcholine, and epinephrine signaling pathways, which affect behavioral, emotional, and physical states and have been associated to psychosis, anxiety, pain, and inflammation(2)(9).The neuroactive ligand receptor interaction pathway was found to be more important during T2 and T3. These pathways correspond to meditation’s claimed benefits on heart rate, stress response, blood pressure, respiratory function improvement, cognitive enhancement, and immune function enhancement (10). Certain pathways, nearly entirely related with T3, are linked to GABAergic synapse, nicotine, and morphine addiction. Our findings are consistent with previous research that links inflammation, immunological response, and neuroactive receptor signaling pathways, providing alternative options for intervention in diseases such as Parkinson’s and multiple sclerosis(11).The absence of medication signatures in the post-meditation phase (T4) emphasizes the significance of consistent practice to preserve therapeutic benefits. The high overlap with drug signatures raises the possibility that people who are taking antidepressants and antipsychotics, may have unwanted side effects from meditation. This is consistent with previous reports of adverse consequences, which include physiological, psychological, psychopathological, and spiritual occurrences(12). We also found signatures with medicines used in hypnotherapy, such as scopolamine, and ones that produce euphoria. During meditation, suppressed memories of traumas, as well as accompanying emotional feelings, are occasionally reported(13). Healthcare professionals who use meditation as an additional therapy should be aware of the possible problems and contraindications(14). Worthwhile to mention, meditation methods were typically taught in a tailored manner by experienced practitioners in ancient times. In contrast, the contemporary era’s internet platforms have facilitated the spread of meditation techniques without cognizance of these practices or intellectual grounding. Our research implies that traditional systems may have evolved to minimize negative consequences and increase benefits. The study also highlights the variability in meditation response and emphasizes the need for a personalized approach to achieve optimum benefits. However, these insights from a specific meditation approach needs to be to further tested in other practices to make the inferences more generalizable. Moreover, gene expression in conjunction with physiological and baseline genomic variations may provide better personalization recommendations. This study revealed unexpected connections between meditation-induced gene profiles and medication reactions, that could provide evidence based application of meditation in clinical settings.

## 3. Materials and Methods

### A. Study Cohort and Data Collection

The study was carried out on 389 transcriptomes (GSE174083), from 106 individuals of a meditation retreat (ClinicalTrials.gov identifier: NCT04366544). The RNAseq data were before the retreat (T1),after an 8-week preparatory phase (T2), 8 days after the meditation retreat (T3), and 3 months after the retreat (T4). Trimmomatic (version 0.39) with default parameters was used on raw reads for adapter removal and trimming(15) followed by STAR for mapping the RNA-Seq data, and DESeq2 (1.40.2) for identification of differentially expressed genes (log2fold changes 1 and pvalue 0.05). To explore inter sample variability, the raw count of samples from T2 and T3 timepoints were TMM normalised prior to DESeq2 (1.40.2) and clusters were identified through hierarchical clustering (Scipy version 1.11.3) and Principal Component Analysis (Sklearn version 1.3.2).

### B. Connectivity Map Analysis to identify interventionable points

The ranked lists of differentially expressed up and down regulated genes in pairwise comparisons of T2vsT3, T2vsT4, and T1vsT2 were used to query the library of gene expression profiles from connectivity map (CMap) data base. Signatures with a connectivity score of *≥* 98 were considered. Gene Ontology analysis was performed on the targets to uncover the enriched pathways and targets influenced by meditation-related gene signatures. Specifically, for inferring pathways and targets similar to those affected by drugs, the enrichment was performed using gene sets exhibiting positive connectivity with drugs and knockdown genes. Conversely, pathways aligning with drugs showing negative connectivity or corresponding to overexpressed genes were deduced as pathways exhibiting opposite effects.

## Data Availability

All data produced in the present study are contained on the manuscript.

## ACKNOWLEDGMENTS

MM and AJ acknowledge financial support from MOA(Ministry Of AYUSH) for Center of Excellence “AyurTech”(S/MOA/MTM/AA/20210105), IIT Jodhpur.Authors acknowledge Debarka Sengupta, IIIT Delhi, for his critical inputs in analysis.

## C. Supplementary Information

All study data are included in the article and/or .SI Appendix.

## Notes

### Competing Interest Statement

The authors have declared no competing interest.

### Clinical Trial

ClinicalTrials.gov identifier: NCT04366544.

### Funding Statement

The fellowship and infrastructure support for the PI and personnel involved in the study was funded by Ministry of AYUSH. The study per se is meta analysis and did not require any funds. This is acknowledged.

### Author Declarations

The study was carried out on 389 transcriptomes (GSE174083), from 106 individuals of a meditation retreat (ClinicalTrials.gov identifier: NCT04366544.

## References

1. M McGee, Meditation and psychiatry. Psychiatry 5 (2008).

2. AB Newberg, J Iversen, The neural basis of the complex mental task of meditation: Neurotransmitter and neurochemical considerations. Med. Hypotheses 61 (2003).

3. S Sadhasivam, et al., Isha yoga practices and participation in samyama program are associated with reduced hba1c and systemic inflammation, improved lipid profile, and short-term and sustained improvement in mental health: A prospective observational study of meditators. Front. Psychol. 12 (2021).

4. S H., Meditation: Process and effects. Ayu 36 (2015).

5. V Chandran, et al., Large-scale genomic study reveals robust activation of the immune system following advanced inner engineering meditation retreat. Proc. Natl. Acad. Sci. 118 (2021).

6. A Subramanian, et al., A next generation connectivity map: L1000 platform and the first 1,000,000 profiles. Cell 171 (2017).

7. M Haider, et al., Anti-sars-cov-2 potential of cissampelos pareira l. identified by connectivity map-based analysis and in vitro studies. BMC Complementary Medicine Ther. 22 (2022).

8. RV Vishnubhotla, et al., Advanced meditation alters resting-state brain network connectivity correlating with improved mindfulness. Front. Psychol. 18 (2021).

9. D Krishnakumar, MR Hamblin, S Lakshmanan, Meditation and yoga can modulate brain mechanisms that affect behavior and anxiety-a modern scientific perspective. Anc. Sci. 2 (2015).

10. AT Tuem KB, Neuroactive steroids: Receptor interactions and responses. Front. Neurol. 8 (2017).

11. R Pacheco, F Contreras, M Zouali, The dopaminergic system in autoimmune diseases. Front. Immunol. 5 (2014).

12. A Cebolla, M Demarzo, P Martins, J Soler, J Garcia-Campayo, Unwanted effects: Is there a negative side of meditation? a multicentre survey. PLoS ONE 12 (2017).

13. JJ Miller, The unveiling of traumatic memories and emotions through mindfulness and concentration meditation: Clinical implications and three case reports. J. Transpers. Psycology 25 (1993).

14. S Joshi, A Manandhar, P Sharma, Meditation-induced psychosis: Trigger and recurrence. Case Reports Psychiatry 2021 (2021).

15. AM Bolger, M Lohse, B Usadel, Trimmomatic: a flexible trimmer for Illumina sequence data. Bioinformatics 30 (2014).

