## Supplementary figures and images for "Signatures of medications in meditation: A Connectivity map analysis of transcriptomes from an inner engineering retreat"

### Supplementary-data_Figure1 copy.jpeg

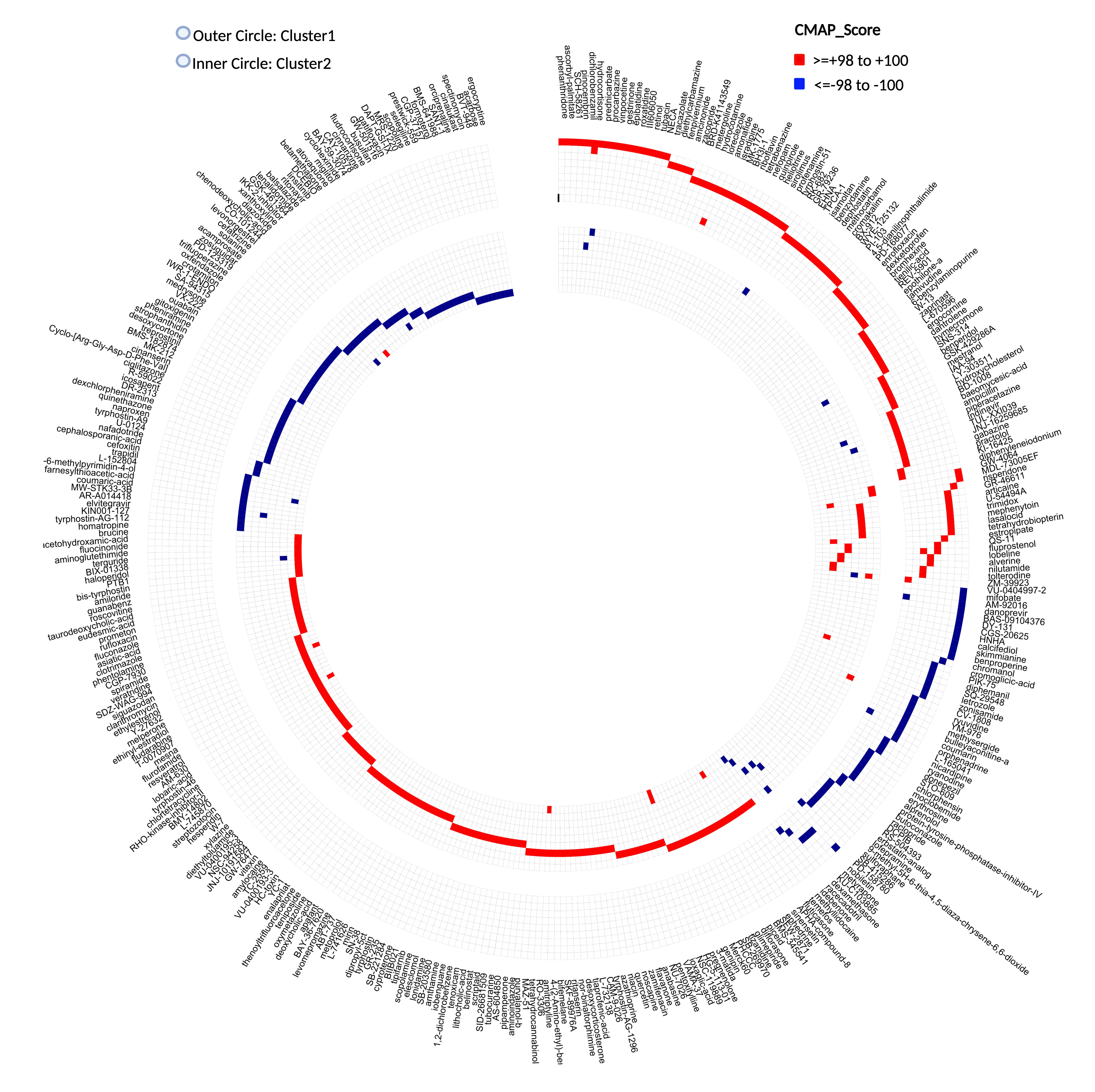
